# Optimizing Clinical Outcome and Toxicity in Lung Cancer Using a Genomic Marker of Radiosensitivity

**DOI:** 10.1101/2020.01.09.20017046

**Authors:** Jacob G. Scott, Geoff Sedor, Michael W. Kattan, Jeffrey Peacock, G. Daniel Grass, Eric A. Mellon, Ram Thapa, Michael Schell, Anthony Waller, Sean Poppen, George Andl, Steven A. Eschrich, Thomas J. Dilling, William S. Dalton, Louis B. Harrison, Tim Fox, Javier F. Torres-Roca

## Abstract

Cancer sequencing efforts have demonstrated that cancer is the most complex and heterogeneous disease that affects humans. However, radiation therapy, one of the most common cancer treatments, is prescribed based on an empiric one-size-fits all approach. We propose that the field of radiation oncology is operating under an outdated null hypothesis: that all patients are biologically similar and should uniformly respond to the same dose of radiation. We have previously developed the Genomic Adjusted Radiation Dose (GARD), a method which accounts for biological heterogeneity and can be utilized to predict optimal RT dose for an individual patient. In this article, we utilize GARD to characterize the biological imprecision of one-size-fits-all RT dosing schemes which result in both over- and under-dosing for the majority of patients treated with RT. To elucidate this inefficiency, and therefore the opportunity for improvement using a personalized dosing scheme, we develop a patient-specific competing hazards-style mathematical model combining the canonical equations for tumor control (TCP) and normal tissue complication probabilities (NTCP). This model simultaneously optimizes tumor control and toxicity by personalizing RT dose using patient-specific genomics. Using data from two prospectively collected cohorts of patients with non-small-cell lung cancer, we validate the competing hazards model by demonstrating that it predicts the results of RTOG 0617. We show how 0617 failure can be explained by the biological imprecision of empiric uniform dose escalation which results in 80% of patients being over-exposed to normal tissue toxicity without potential tumor control benefit. In conclusion, our data reveals a tapestry of radiosensitivity heterogeneity, provides a biological framework that explains the failure of empiric RT dose escalation, and quantifies the opportunity to improve clinical outcomes in lung cancer by incorporating genomics into RT.

## Introduction

The empiric basis of radiation therapy (RT), the most commonly utilized therapeutic agent in clinical oncology, has gone unmodified for over 70 years. RT is prescribed based on a uniform, one-size fits all approach, delivering small daily doses of RT over several weeks (i.e. fractionation). This fractionation approach is based on studies performed in rams and rabbits by Regaud, Schinz and Slotopolsky over 100 years ago^1–5^ – and the standard total doses for control of sub-clinical, microscopic and macroscopic disease (50, 60 and 70 Gy) were established in the 1960s based on tumor control probability models for head and neck cancer patients^6,7^. Although there has been a recent interest in hypofractionation, all of these schedules have also been empirically derived.

Although RT remains a critical curative agent for cancer, it has yet to adapt a biological basis in the clinic. That radiation response is heterogeneous, even within disease groups, is well known. Further, that this heterogeneity is driven and influenced by changes in the tumor genome is now accepted – indeed, large-scale classification efforts have been performed to understand these differences.^8–10^ Additionally, there have been several efforts to understand surrogate genomic metrics for individual patient’s resistance to radiation^**?**^, as well as imaging-based studies^11^. We previously proposed that the gene expression-based radiosensitivity index (RSI), a surrogate for intrinsic cellular radiosensitivity, and the genomic-adjusted radiation dose (GARD), an individualized quantitative metric of the biologic effect of RT, could serve as the first approach to biology-based RT. Both RSI and GARD have been validated in multiple clinical cohorts and disease sites as a predictor of clinical outcome in patients treated with RT^12–16,16–20^. Importantly, the Lancet Oncology commission identified GARD as an candidate biomarker for personalized radiation oncology^21^, and the EORTC identified the totality of the evidence surrounding RSI/GARD as a priority for phase III trials^22^. In addition, two recent independent studies from Lund University and Milan provide corroborative evidence that RSI is predictive of RT benefit in breast and head and neck cancer; a predictive biomarker^23^.

We hypothesize that given the known heterogeneity of cancer, there is an optimal RT dose for each patient that maximizes tumor control and limits toxicity; an ideal personalized therapeutic ratio. Further, we hypothesize that this optimal dose is at least partly defined by tumor biology, as has been posited before^24^. In this manuscript, we utilize RSI/GARD to calculate the optimal RT dose for each patient in a cohort of 1,747 NSCLC patients. We identify three distinct radiobiological cohorts in NSCLC: (i) Sensitive patients who are biologically optimized at current standard of care RT dose, (ii) Intermediate patients who may benefit from moderate genomically-directed RT dose escalation and (iii) Resistant patients who require RT dose beyond standard of care. To further understand the consequences for outcome in each of these groups, we develop a novel mathematical model for outcome. This model combines our genomic approach to calculate optimal dose and canonical models of tumor control and normal tissue complications to calculate a patient specific predicted outcome which includes morbidity and mortality from specified causes. To make this combined model simpler to use clinically, we further present decision-support software that provides the first approach to biologically optimize clinical outcome and toxicity for each individual patient. Our data reveals a tapestry of radiosensitivity heterogeneity, provides a biological framework that explains the failure of empiric RT dose escalation, and quantifies the opportunity to improve clinical outcomes in lung cancer by incorporating genomics into RT.

## 1 Materials and Methods

### 1.1 Patients

We utilized patients from Total Cancer Care (TCC), a prospective IRB-approved data and tissue collection protocol active at Moffitt and 18 other institutions since 2006^25^. Tumors from patients enrolled in TCC protocol were arrayed on Affymetrix Hu-RSTA-2a520709 (Affymetrix, Santa Clara, CA), which contains approximately 60, 000 probesets representing 25, 000 genes. Chips were normalized using iterative rank-order normalization (IRON)^26^. Batch-effects were reduced using partial-least squares (PLS). The normalized, de-batched expression values for 1,747 NSCNC (NSCLC) samples and the ten RSI-genes were extracted from the TCC database. To quantify the impact of the optimal dose on tumor control and toxicity, we utilized a subset of 60 patients with Stage 3 NSCLC treated at Moffitt with post-operative RT, which has also been previously described. The clinical endpoint was local control. The median follow up (based on the reverse Kaplan-Meier method) in censored patients free from local failure was 59.5 months (95 % CI:38.0-68.5 months)^27^.

### 1.2 Radiosensitivity Index (RSI)

RSI scores were previously generated^18^. RSI was previously trained in 48 cancer cell lines to predict cellular radiosensitivity as determined by survival fraction at 2 Gy (SF2)^17^. Each of ten genes in the algorithm is ranked based on gene expression (highest expressed gene is ranked at 10 and lowest at 1) and RSI is calculated using the pre-determined equation:

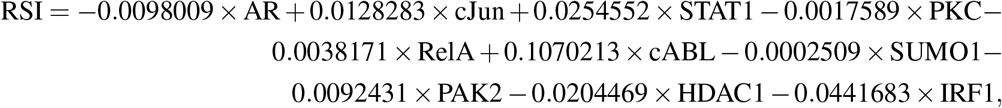

which has been presented previously^17^.

### 1.3 Genomic Adjusted Radiation Dose (GARD)

GARD, a unitless measure of genomic radiation effect, has been previously described^18^. Briefly, it is derived using the LQ model (*S* = *e*^*−nd*(*α*+*βd*)^), and the individual RSI and the radiation dose/fractionation schedule for each patient. First, a patient-specific (genomic) *α*_*g*_ is derived by substituting RSI for Survival (S) in LQ equation, yielding:

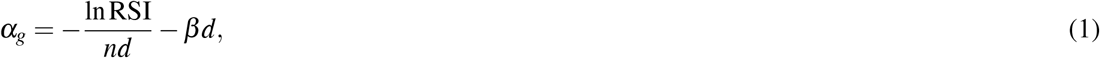

where dose (d) is 2Gy, n is the number of fractions (here *n* = 1, recall as we are deriving *α*_*g*_ from RSI which was trained on an SF2 assay), and *β* is a constant 0.05*/*Gy^2^. GARD is then calculated using the classic equation for biologic effect, GARD = *nd*(*α* + *βd*), the patient-specific *α*_*g*_ calculated as per equation 1, and the number of clinical fractions (*n*_*c*_) and dose per fraction (*d*) received by each patient. It is worth noting that in the case when a patient receives a single 2Gy fraction, the *βd* terms drop out, and this genomic 2Gy equivalent (GARD_2*Gy*_, similar in spirit to EQD2) is

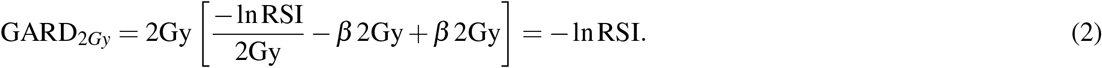

For a clinical GARD (which we will from here on denote GARD_*c*_, one can simply scale this by the number of fractions given (assuming 2Gy fraction) yielding 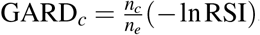, where *n*_*i*_ where *i ∈* {*c;e*}, is the number of doses given in the *c*linical scenario (individualized per patient), and the *e*xperimental (by definition *n*_*e*_ = 1) conditions. However, for generality we define the *c*linical GARD as

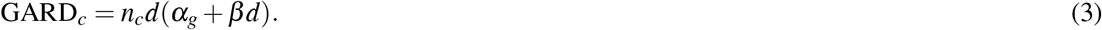

A GARD cut-point of 33 was previously identified and published for the lung clinical cohort^18^, and will be utilized going forward in this manuscript. It is worth noting, however, that this cutpoint will differ for each cohort. Briefly, this threshold is found by minimizing the *p −* value for the Kaplan-Meier statistic by iterating through possible cutpoints for GARD in a cohort with known outcomes. For the cohort used in this manuscript, for which we identified a GARD_*T*_ = 33, we show this analysis in supplemental section 3 (Step by step derivation of RxRSI), and include the code in the linked github repository.

### 1.4 Biologically-Optimized Personalized RT dose (RxRSI)

Here, we define a new term, RxRSI, as the physical dose (in Gy) required to achieve a previously identified GARD threshold (GARD_*T*_, in this paper, GARD_*T*_ = 33) in a cohort of lung cancer patients treated with post-operative RT^18^. RxRSI is calculated using the following formula:

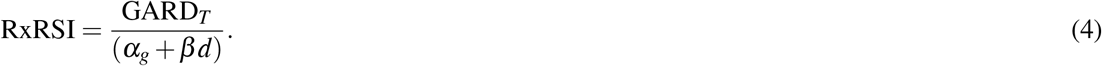

Where *α*_*g*_ is calculated based on each individual patient’s RSI per equation 1 and *β* is a constant (0.05*/*Gy^2^). When comparing RxRSI to the empiric dose received by patients in the lung cancer clinical cohort, we defined that the RxRSI and empiric dose matched if they were within 10% of one another. As GARD was developed based on standard fractionation, we further assume that RxRSI is delivered in a similar manner (i.e. dose per fraction is *∼*2Gy). Methods for calculating optimal doses for altered fractionation schedules can be calculated using the same method, but *β* needs to be estimated in a different manner.

### 1.5 Genomic Radiation Treatment Planning

To quantify the impact of the biological optimal RT dose on outcome and toxicity we integrated the algorithms and equations that define RSI, GARD and RxRSI into radiation treatment planning software. We generated 30 RT plans to match the anatomical and biological diversity in the 60 patient cohort. Plans were created for the following biological conditions (RxRSI = 48 Gy, RxRSI = 54 Gy, RxRSI = 62 Gy, RxRSI = 74 Gy, RxRSI = 88 Gy and RxRSI = 95 Gy)(Supplementary Figure 1). Dosimetric parameters for normal tissue including mean heart dose, mean esophagus dose, and mean right and left lung dose, were calculated for all genomic plans. We utilized the resulting data to generate a linear model to estimate the impact of dose personalization on normal tissue (see Supplemental Figure 2).

### 1.6 Linear Model for Normal Tissue Estimates

The mean dose to each normal tissue target (heart, left lung, right lung and esophagus) were calculated across the 30 genomic plans developed. Mean normal tissue dose was plotted against PTV prescription dose to obtain a Pearson’s correlation coefficient for mean heart, left lung, right lung, and esophageal dose (*R*^2^: 0.98, 0.99, 0.97, 0.99, respectively). These linear relationships were then used to calculate an approximate mean dose to normal tissue on a Gy^*−*1^ basis.

### 1.7 Normal Tissue Toxicity

To create a combined model of tumor control and NTCP, we required a model of excess toxicity probability for each orgran at risk (OAR) per Gy delivered. Calculations for relative risk for a given dose received or dose adjustment was accomplished using different methods for each tissue site, depending on the available data and recommendations in the literature. When possible, data on rate of complication per dose received was used, or a quantitative NTCP model which has the benefit of flexibility in choosing dosing parameters. For generalizability, specific dose-toxicity endpoints were not referenced. This method can be extended to any OARs, but for this manuscript we focus on the three main drivers of complications in NSCLC radiation therapy that have quantifiable models: pneumonitis, esophagitis and radiation induced heart disease.

In the QUANTEC review of lung complications, the primary endpoint is radiation pneumonitis^28^. The reviewers conducted a meta-analysis of applicable studies and performed logistic regression on rates of radiation pneumonitis versus mean lung dose (MLD),

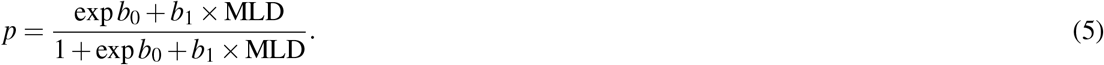

Parameters for *b*_0_ and *b*_1_ were calculated for a model in the above form. The QUANTEC reported recommendations for toxicity endpoints for the esophagus were inconclusive due to the volume-dependent effect of the available data^29^. Two of the studies, both published in 2005, provided quantitative models in the form of the Lyman-Kutcher-Burman equation, with parameters *m* and *TD*50 that were within bounds of the confidence intervals^30,31^,

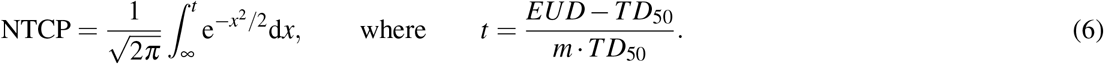

Cardiac complications due to radiation were modeled as a fixed rate of 7.4% increased risk per 1 Gy dose received by the heart. The endpoint included coronary events as defined by myocardial infarction, coronary revascularization, or death from ischemic heart disease^32^.

### 1.8 Statistical Methods

A survival regression model was used to quantify the impact of individual GARD on local control. By applying the cut-point of 33 to define two strata, a time-dependent parametric model was developed that could then be adjusted by normal-tissue effects. This initial calculation of parameters for a Weibull distribution was done using the Surv-Reg package in R, of the form *H*(*t*) = *λt*^*ρ*^, as the hazard function, and 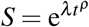 as the survival function. The adjusted outcome models were developed in python in the defined form such that

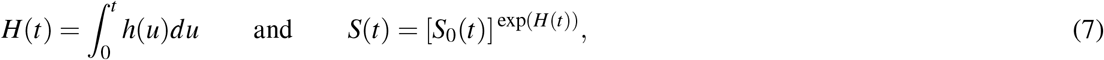

and implemented in the Dash open source library for data visualization.

## 2 Results

### 2.1 The biological optimized dose (RxRSI) identifies three distinct radiobiological clinical cohorts in NSCLC

As we have demonstrated before^18^, there is wide heterogeneity in the radiation sensitivity in NSCLC. In Figure 1A, we plot the distribution of of RSI in a large (1,747 patients) cohort of NSCLC patients from the TCC cohort (range: 0.079-0.752). Of note, there is a bimodal distribution of RSI dose across this population, suggesting that a uniform, one-size fits all approach to RT dose is sub-optimal for the majority of patients. Taking into consideration a separate cohort of post-operative NSCLC patients with known clinical history, including post-operative radiation dose, we calculate RxRSI (the physical dose predicted to optimize biological outcome). The distribution of this calculation across this cohort (Figure 1B, also see supplemental figure S5 for the raw RSI:*α*_*g*_ transformation for this cohort), reveals three distinct radiobiological cohorts in this subset of patients: 1. Radiosensitive patients which achieve RxRSI at current standard of care RT dose (50 Gy or less in this post-operative setting), 2. Intermediate sensitivity which achieve RxRSI within the standard of care accepted range(50-70Gy) and 3. a radioresistant group which require doses above standard of care (>70 Gy for post-operative RT) to achieve RxRSI.

**Figure 1.**
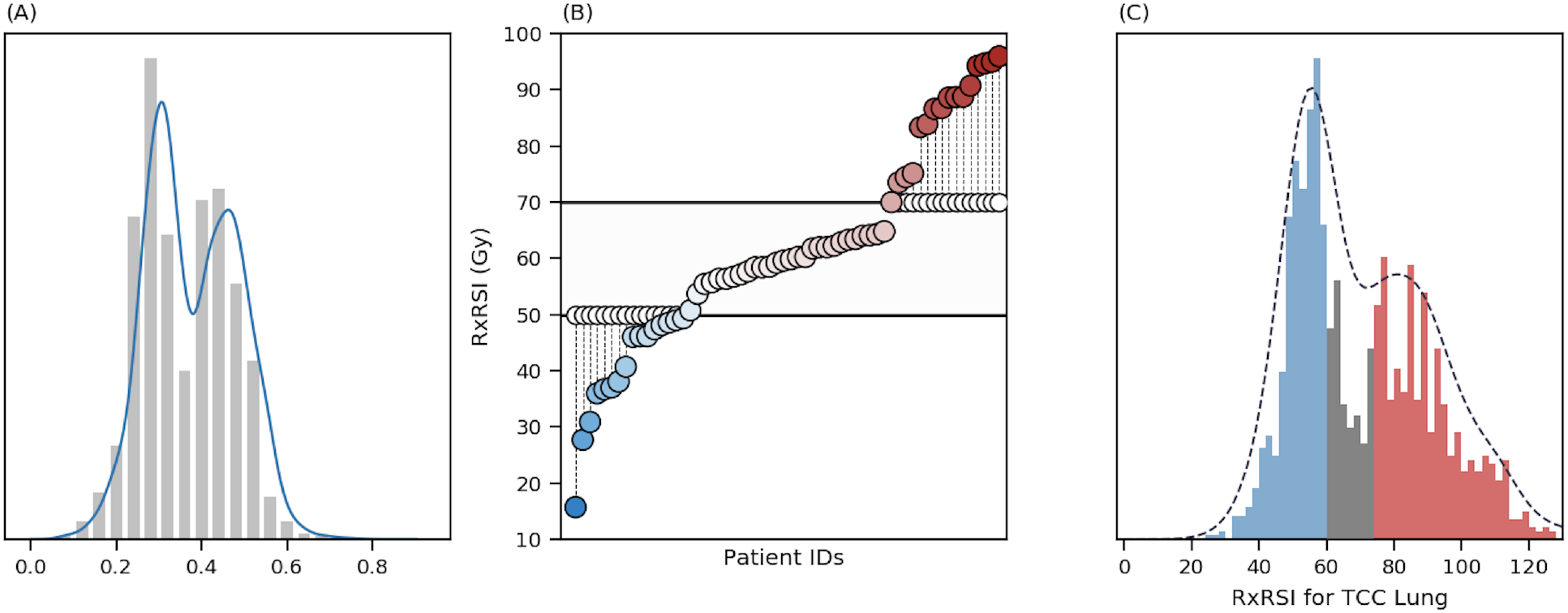
We identify three distinct groups of patients from genomics and standard RT dosing schedules. **(A)** Distribution of RSI in a cohort of 1,747 patients with NSCLC in the TCC cohort. **(B)** Calculating RxRSI (the physical dose required to achieve an optimized biological outcome) for each patient in a clinical cohort of 60 patients with known clinical outcome, dose received and RSI reveals three groups: patients who require less than SOC dose (50Gy), patients who require a dose within the SOC range (50-70Gy) and patients who require more than the SOC dose (*>* 70Gy). **(C)** Translating to primary radiation doses and a larger (TCC) cohort, we see that there is a subset of patients who are optimized by 60Gy (blue), a small subset of patients would benefit from moderate (up to 74Gy - grey) and a large cohort (red) who would need greater than 74Gy.

**Figure 2.**
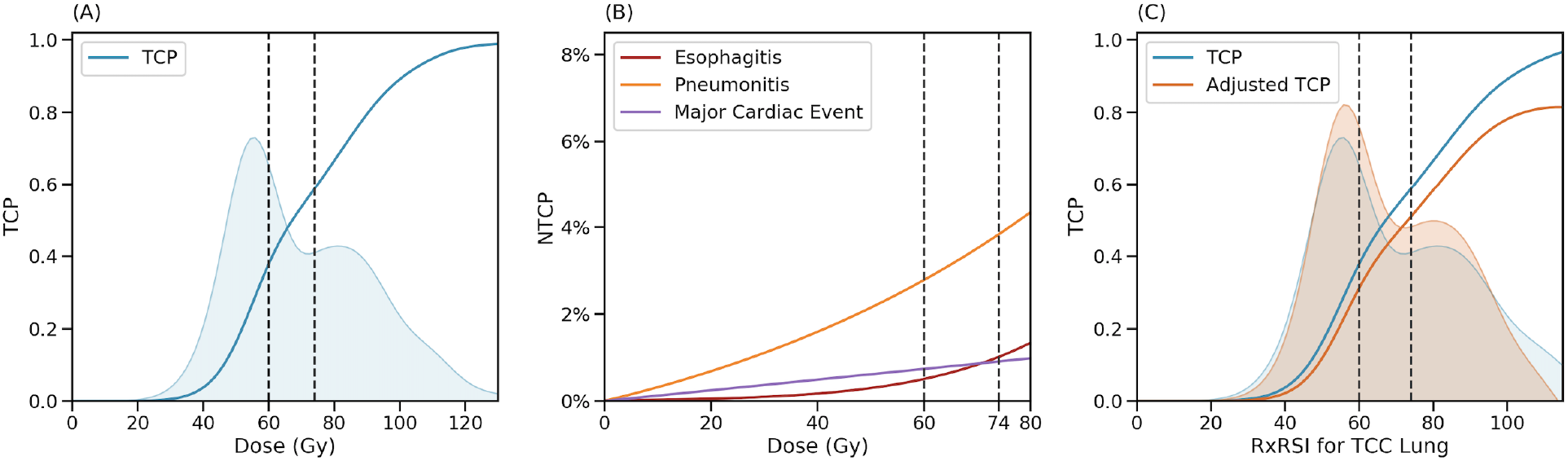
Combined TCP and NTCP model). **(A)** The cumulative distribution function of the (bi-modal) RxRSI is a TCP curve (and approximates a sigmoid). **(B)** Probability of grade 3 or greater toxicity with dose for each of esophagus (purple), lung (yellow) and heart (blue). **(C)** TCP (blue) corrected by NTCP (yellow) as a function of dose.

Translating this calculation to the larger (TCC) cohort, and into primary radiation dosing, we see a similar split into three groups, but now notice another interesting finding: in the area of recent dose-escalation (60-74Gy) there is a very low number of patients, suggesting that very little is to be gained in this region. Figure 1C shows three regions, (blue) where any patient would be optimized by 60Gy, (grey) where moderate dose escalation to 74Gy is required and (red) a large region where doses above 74Gy would be required (of note, this region contains approximately 42% of the population, which happens to be approximately the percentage who experience local failure with chemoradiation).

### 2.2 Empiric RT dose is biologically imprecise and results in an inefficient distribution of RT-related toxicity and clinical benefit

Historical models of radiation response have always considered either tumor control or normal tissue complications. This was all that was possible, because no estimate of *required* dose was available. With the advent of our predicted optimal dose, we now have the ability to quantify *excess* dose received by individual patients when receiving empiric dosing. To quantify the untoward effects of empiric RT dose then, we generated 30 radiation treatment plans representing the distribution observed for RxRSI in the lung cancer patients treated with post-operative RT. We calculated the excess normal tissue dose delivered (when the patient was given more than RxRSI) or the additional normal tissue dose required (when patients receive a dose lower than the RxRSI). For 25% of the patients in our cohort, the empiric dose and RxRSI matched while for 75% it did not match (supplementary table 1-4). We then calculated the impact on normal tissue dose and toxicity of actually delivering RxRSI for each patient using RT doses within standard of care guidelines (RT dose 50-70 Gy). In sensitive post-operative patients, adjustment to the RxRSI (set to a minimum dose of 50 Gy) would have resulted in an overall mean dose **decrease** to the esophagus, right and left lung and heart (supplementary table 4). In intermediate post-operative patients, adjustment to the RxRSI would also have resulted in a mean **increase** in dose to normal tissue (supplementary table 2). The mean increase in normal tissue dose for intermediate patients (RxRSI>Dose received) is very similar to mean decreases experienced by sensitive patients (RxRSI<Dose received). Thus, since resistant patients are not adjusted because RxRSI is above standard of care (supplementary table 3), the overall risk profile for normal tissue complications for the whole population is not expected to be affected by the dose adjustments proposed by RxRSI. The predicted impact of personalized dose adjustments on normal tissue toxicity is shown in supplementary table 5. In summary our data demonstrates that it is possible to deliver RxRSI to 75% of the patients without changing the overall toxicity profile for the whole population.

### 2.3 Development of combined mathematical model to correct tumor control by toxicity from excess dose

To estimate the clinical potential for personalized prescription RT dose beyond simply tumor control, we developed a mathematical model to utilize genomic markers of radiosensitivity to optimize radiation outcomes considering **both tumor control and individual toxicity**. While great efforts have been made to understand the untoward effects of radiation over the decades^33^, **knowledge of excess dose for an individual patient has not been possible**, and this information has not been able to be incorporated into personalized predictions. Our genomic framework, in particular the estimate of required dose, RxRSI, provides a first estimate of this. To understand the combined contributions of tumor and excess normal tissue effects on outcomes, we have created a competing hazards style risk model. We term the outcome the “penalized local control” (pLC), which includes local recurrence (akin to the classic Tumor Control Probability]^34,35^) and events related to RT-related toxicity (styled after NTCP), but does not account for death due to disease progression or other causes. The penalized local control curve for a population is calculated as *S*(*t*) = *C*_1_*S*_1_(*t*) + *C*_2_*S*_2_(*t*) where *C*_*i*_ (*i ∈* 1, 2) represents the fraction of patients that receive a tumor dose that is either adequate (*i* = 1, RT dose *≥* RxRSI), or inadequate (*i* = 2, RT dose *<* RxRSI), and *S*(*t*) is the survival function derived from the individual cohort’s KM analysis (see Methods 1.8). Finally the convolved survival curves is then adjusted for the predicted toxicity hazard ratios as per Methods 1.7:

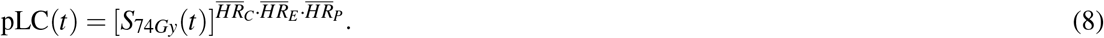

Here 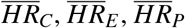 are the risks for each adverse outcome including cardiac, esophagitis and pneumonitis, respectively).

### 2.4 Combined tumor control and NTCP model accurately predicts the outcome of RTOG 0617

To validate the combined TCP and NTCP model, we designed an *in silico* clinical trial (a phase *i* trial if you will^36^), to match the recent trial of uniform dose escalation in NSCLC (RTOG 0617 60Gy vs 74Gy). As in RTOG 0617 we assigned (uniformly at random), 200 patients for a 74Gy arm, and 200 patients for a 60Gy arm. We calculated the expected clinical outcome for each arm based on an estimate of tumor control and toxicity for each patient. We performed 100 iterations of this *in silico* trial, randomly assigning an RSI value to each in silico patient using data from the TCC NSCLC cohort. This trial is schematized in Figure 3A. Of note, these two empiric distributions (Clinical and TCC) are striking similar to one another, sharing two dominant modes with 1% of one another (see Supplementary Figure 4).

**Figure 3.**
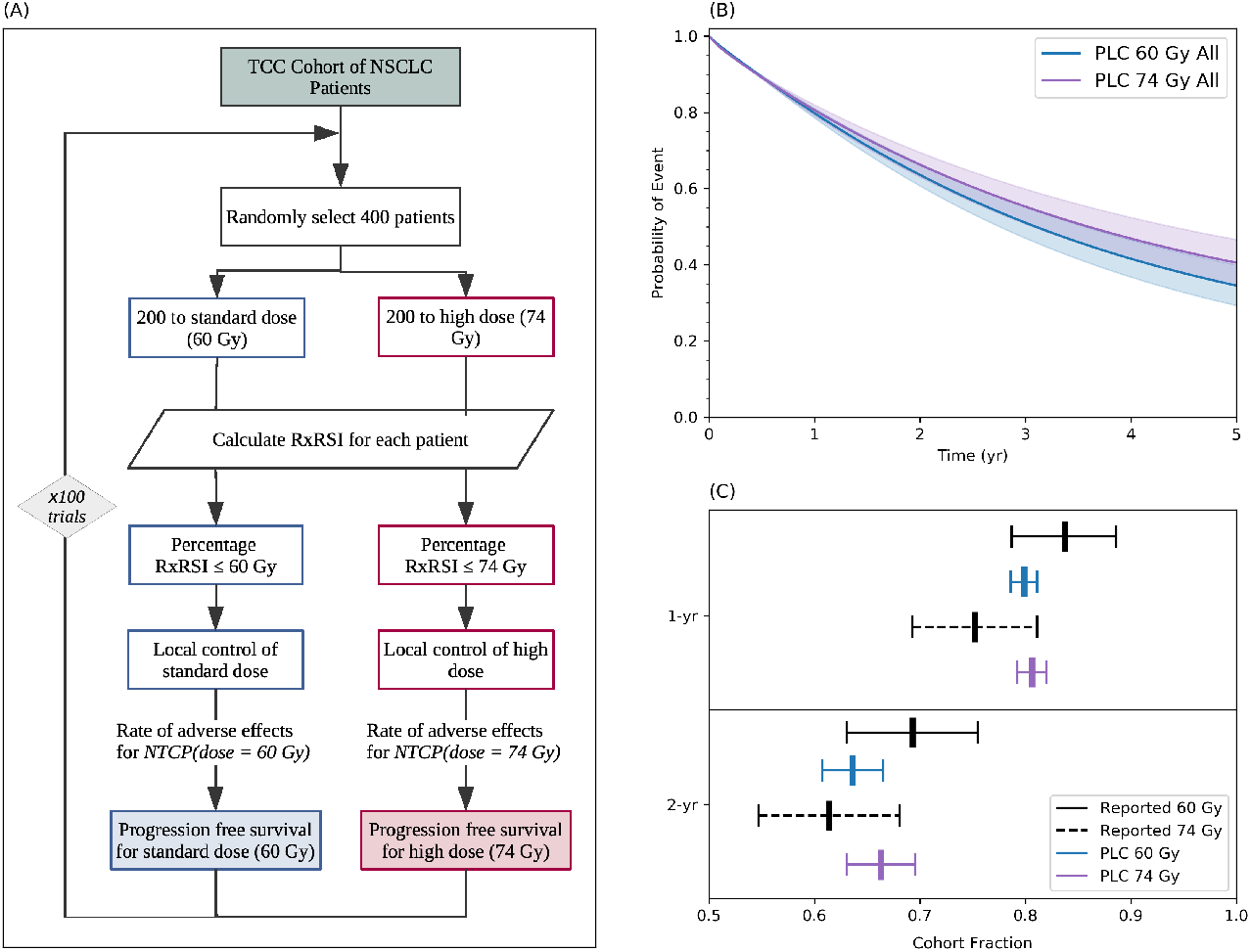
An *in silico* trial of dose escalation using the competing outcomes model in NSCLC matches the outcomes of a recent cooperative group trial. **(A)** Schematic of our *in silico* trial designed to match RTOG 0617, with patients drawn uniformly at random from the TCC cohort. **(B)** A Kaplan-Meier curve depicting penalised local control (pLC). The 60 and 74Gy arms are predicted to have statistically indistinguishable outcomes (penalized local control) through 5 years. **(C)** Using the combined model accurately predicts the results of RTOG 0617.

As shown in Figure 3B, the combined model predicts that uniform dose escalation to 74 Gy to unselected patients would result in no radiation-associated overall gains when compared to 60 Gy, consistent with the results observed in the actual clinical trial. In addition, as shown in Figure 3C, the model correctly predicts the 1 and 2 year local control observed in RTOG 0617. To further understand the biological underpinnings to explain this result, we determined the proportion of patients that were expected to derive a benefit from dose escalation to 74 Gy. As seen in Figure 4A (blue group), 39.6% of the patients achieved or exceed RxRSI at 60 Gy. Only an additional 18.6% reached RxRSI at 74 Gy (grey group). However, our model predicts that still about 41.7% of the patients may need higher doses (>74Gy, red group). Thus, in an unselected population, uniform dose escalation to 74 Gy benefits only a minority (grey only) of patients and exposes the majority (blue and grey) of patients to additional toxicity, obfuscating any radiation-associated clinical gains. However, a targeted dose escalation strategy, where only patients in the cohort of patients with intermediate radiosensitivty (RxRSI, 62-74 Gy) receive 74 Gy would be expected to improve the local control for the whole cohort by an absolute 4.1% at 2 years, and 7.8% at 5 years (Figure 4B). This small, but significant gain, would increase to 4.3% at 2 years and 8.1% at 5 years if patients were given the exact dose that was predicted to optimize their outcome, rather than the full 74Gy, as they would be spared the additional toxicity. Taken to the logical limit, where each patient is given only the dose they need, to an upper limit of 80Gy (lower bound of 45Gy), would further increase the outcomes by another 2.8% and 5.4% at 2 and 5 years, respectively, highlighting the opportunity when radiation therapy is truly personalized.

**Figure 4.**
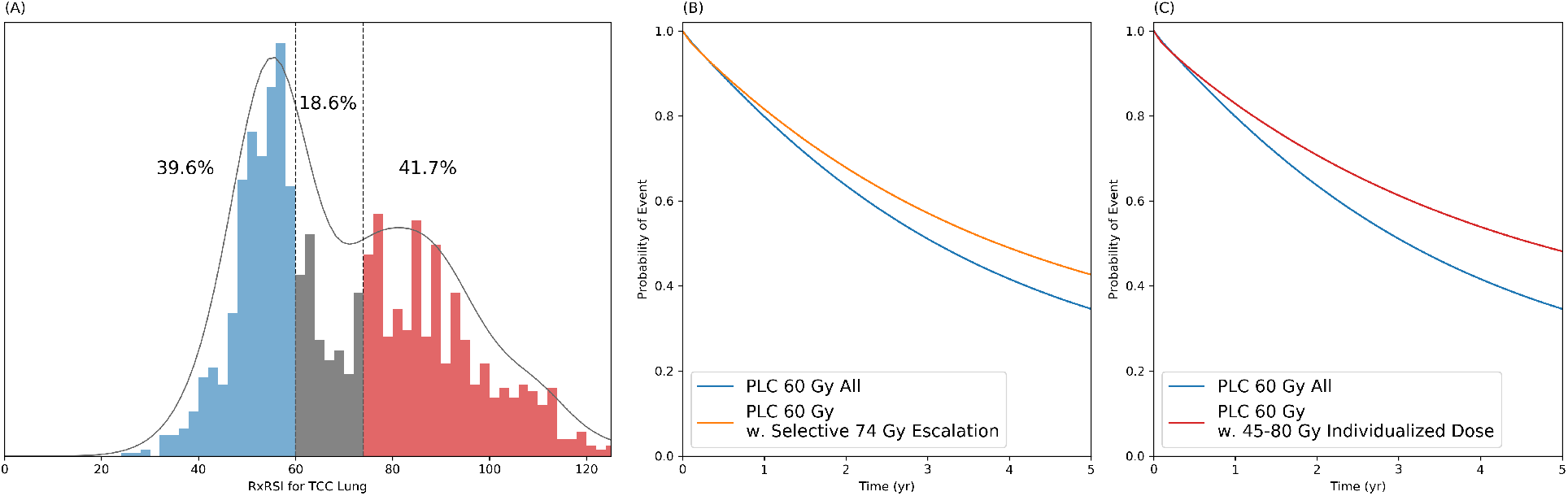
Empiric dose escalation reached a local optima at 60Gy, but personalized dosing offers significant benefits with current technology. **(A)** A radiation dose of 60Gy will provide optimal tumor control for approximately 40% of the population, and escalation to 74Gy will only optimize a further 18.6%, while exposing all to additional toxicity. **(B)** A Kaplan-Meier curve depicting an *in silico* trial of 60Gy vs. 74Gy, with escalation only for those the RSI based model predicts who would benefit. **(C)** A Kaplan-Meier curve depicting an *in silico* trial of predicted optimal dose in the range 45-80Gy.

## 3 Discussion

In this paper, we present a clinically-feasible system to personalize RT prescription based on biological parameters, and quantify the clinical opportunity for improved clinical outcomes inherent in personalized RT for patients with NSCLC. Our proposal for personalized RT prescription is based on three parameters: (i) RSI which defines the patient’s individual tumor radiosensitivity; (ii) GARD, which defines the individualized clinical effect of a given dose of RT in a given patient with a distinct RSI; and, (iii) RxRSI, the biologically-optimal RT prescription dose, which we define as the prescription dose required to achieve a GARD target value associated with improved clinical outcome. Personalized RT prescription provides an alternative to the empiric-based one-size fits all approach that is currently standard in the field.

In two separate cohorts of patients with NSCLC, with genomics derived from surgical specimens, we showed that prescribing uniform, empiric-based RT dose is biologically imprecise, with 75% of patients we analyzed receiving non-optimal doses of RT. Conversely, we showed that the personalized, RxRSI-based prescription approach can deliver optimal doses to up to 75% of the patients in the clinical cohort even when we restrict ourselves to a dose range within the standard of care, and without any further clinical information like tumor size, which would likely improve predictions. This optimal dose can be achieved without an overall increase in expected normal tissue toxicity for the whole cohort. To quantify the clinical potential of personalized RT prescription to improve outcomes in lung cancer, we developed a first-in-class mathematical model combining tumor control and normal tissue toxicity. The model assumes an ideal biological dose to maximize tumor control and estimates outcome based on whether the RxRSI is achieved, then incorporates a penalization scheme based on the added toxicity to which patients are potentially exposed when their RxRSI is exceeded. At this stage in the model development, we have kept NTCP estimates based on population averages, and have attempted to propagate the (large) uncertainty that comes with this abstraction through the data. We have made a first attempt to propagate this error through our mixed modeling framework in the supplements, the general findings for RTOG0617 are strengthened, and the findings for our proposed dose escalalation are somewhat weakened, highlighting the need for personalized planning, not just avatar based OAR calculations. In future, we hope to be able to personalize prediction of NTCP as well, but this lies outside the scope of this work.

To validate the model, we tested it using published data from RTOG-0617, a Phase 3 randomized trial in lung cancer that assessed whether a uniform 14 Gy dose escalation would result in clinical gains in lung cancer. The model correctly predicted both qualitatively and quantitatively the (counter-intuitive) trial outcome: that uniform, empiric dose escalation to 74 Gy does not result in any radiation-associated clinical gains, which it explains is secondary to the potential gains in tumor control being outweighed by the number of patients exposed to additional toxicity. However, the model predicts that a personalized strategy to deliver 74 Gy only to the patient subset most likely to benefit (RxRSI 62-74 Gy) would have improved the radiation-associated outcome for the whole cohort by 7.8% in local control at 5 years. Thus, we propose that the delivery of biologically-inaccurate RT doses results in a significant detriment of clinical outcome for lung cancer patients treated with RT.

While the classic LQ model predicts that every individual in a population has the same opportunity to benefit from uniform dose escalation, the RxRSI model predicts that only a minority of patients (16.2% in this analysis) have the opportunity to benefit from dose escalation to 74 Gy. This opportunity to benefit is outweighed by potential increase toxicity to the rest of the patients. Inspecting the distribution of RSI in the two cohorts for lung cancer also illustrates an interesting point. Dose escalation from 45 - 60Gy results in capturing the lion’s share of the patients in the first peak of the distribution. However, escalation from 60 - 74Gy only captures the tail of the first mode, and does not affect the second peak. This may explain how uniform dose escalation to 60 Gy shows benefit to the entire population, as the benefit outweighs the harm. In addition, our model postulates that 42% of the patients are still undertreated at 74 Gy, which is consistent with the local failure rate reported in RTOG0617^37^. We postulate that the distributions we measured here are conserved, and further analysis of them in different disease sites could provide insight into opportunities for personalized dose escalation and de-escalation. On the strength of this analysis, we submit that our lack of understanding of biological heterogeneity, and how to treat it, explains the failure of biologically naïve uniform RT dose escalation.

The framework to personalize RT prescription presented in this paper has a number of advantages over the current empiric approach. First, it accounts for biological heterogeneity that is specific to RT, updating the naive assumption of homogeneous biology across our patients, which is inherent in the empiric approach. Second, since it uses biological information to formulate an optimized and personalized RT prescription dose, it requires that genomic data be collected for every patient. This provides the framework to identify novel biology that impacts RT benefit. Thus the precision RxRSI model is only the first step towards a more efficient and optimal approach to RT prescription in which toxicity and tumor control can be quantitatively optimized in parallel by maximizing the survival function (equation 8) as part of the same inverse problem we are accustomed to solving for physical dose distributions. In contrast, multiple Phase 3 clinical trials have demonstrated that additional clinical benefit from the empiric approach is unlikely^37–41^. Critically, as we have demonstrated, this novel personalized system can be utilized within the standard of care framework for RT dose, and can be done so without the need for additional equipment or medicines.

While significant interest has been focused on the development of better therapeutic agents including targeted agents and immunotherapy, RT remains a fundamental curative treatment for the majority of patients with cancer. It has been estimated that 40% of all cancer cures are due to RT^42^. In contrast, to date, no targeted agent or immunotherapy has shown similar curative potential in solid tumors. Shifting to a biology-based system will provide a new direction for radiation oncology with multiple opportunities to improve clinical outcome. And that opportunity is not small. Approximately, 70% of all cancer patients receive RT which translates to about 850,000 patients in the US^43^. A moderate improvement in RT-based cures of 5% would represent an additional 42,500 patients potentially achieving cure. According to the American Cancer Society, this is approximately the same number of patients that die from breast cancer every year in the US.

In conclusion, radiation oncology has employed an empiric uniform approach to prescribe RT that is based on models developed and published over 70 years ago. We demonstrate that this one-size fits all approach is biologically inaccurate for the majority of patients, and results in significant detriment of clinical outcome for patients treated with RT. We propose a new paradigm, where the field updates its assumptions by acknowledging the biologically heterogeneity of tumors and moves towards the delivery of biological optimal doses of RT.

## Data Availability

Please see attached spreadsheet.

## Acknowledgements

JGS would like to thank the NIH Loan Repayment Program for their generous support and the Paul Calabresi Career Development Award for Clinical Oncology (NIH K12CA076917). JFTR acknowledges support from the DeBartolo Personalized Institute.

